# Intact corticostriatal and altered subcortical circuits in chronic pain

**DOI:** 10.1101/2021.09.08.21263285

**Authors:** Su Hyoun Park, Anne K. Baker, Vinit Krishna, Katherine T. Martucci

## Abstract

Previous research has demonstrated the importance of the corticostriatal circuit in chronic pain. By focusing on nucleus accumbens (NAcc) circuits related to reward, we aimed to clarify how altered brain reward systems contribute to chronic pain. Using resting-state functional magnetic resonance imaging, we compared NAcc-medial prefrontal cortex (MPFC) functional connectivity in patients with fibromyalgia vs. healthy controls. Among patients, we analyzed the extent to which functional connectivity correlated with clinical measures. We also examined NAcc functional connectivity to subcortical regions. Lastly, we compared our results to a separate dataset of patients with chronic back pain. We identified robust NAcc-MPFC functional connectivity among patients with fibromyalgia and healthy controls, with no significant group differences. We found a positive correlational trend between NAcc-MPFC functional connectivity and total mood disturbance. Notably, patients with fibromyalgia showed significantly reduced functional connectivity of the right NAcc with mesolimbic circuit regions compared to controls. These results were largely similar to the results from the separate dataset. Our results provide novel evidence of intact corticostriatal but altered subcortical functional connectivity of the NAcc during resting-state in chronic pain and suggest that measured connectivity may relate to changes in mood and the level of cognitive demand during fMRI-based measurement.

**Perspective:** This article indicates complex brain valuation system alterations associated with chronic pain. Our findings expand our understanding of the valuation system and its relationship to clinical presentation in patients with fibromyalgia.

## INTRODUCTION

Reward/valuation and motivation is central to the decision-making process and human survival and powerfully modulates the experience of pain (1). Neuroimaging studies have identified a critical role for brain reward/valuation systems in pain processing (2–5). Chronic pain is often comorbid with dysfunctional motivation processes (6–8), and extensive previous studies provide evidence of reward circuitry alternation in chronic pain (5,9–13). Thus, altered mesolimbic reward/valuation systems may reflect altered reward behavior and together be important contributor to chronic pain.

In the human brain, reward processing involves the cortico-basal ganglia network, including the nucleus accumbens (NAcc), and other structures such as the medial prefrontal cortex (MPFC), amygdala, hippocampus, thalamus, and anterior cingulate cortex (ACC) (14–16). Connections between these regions form a complex brain reward network and serve as critical pathways for reward processing (15,17–19). Especially, the NAcc is a key subcortical region within the ventral striatum that is strongly implicated in brain reward processing (14,17). Dopaminergic projections from the midbrain to the NAcc are a key neural component of reward processing (17), and reciprocal connections exist between the NAcc and MPFC.

Prior studies using functional magnetic resonance imaging (fMRI) to assess functional connectivity between the NAcc and brain areas involved in reward have provided evidence of reward system dysfunction in patients with chronic pain (9–11). For example, when patients with chronic back pain receive thermal stimulation and perform coinciding self-report pain-monitoring task, they exhibit greater functional connectivity between the NAcc and MPFC compared to healthy controls (10). Another study revealed that when sub-acute back pain patients performed continuous self-report pain-monitoring task during a resting-state fMRI scan, connectivity between the NAcc and MPFC predicts pain chronification (i.e., greater NAcc-MPFC functional connectivity in patients whose pain persisted vs. patients who recovered) (11). However, to date, no studies have evaluated the NAcc-MPFC circuit in patients with back pain during a resting-state status when no task is involved.

Brain reward system alterations are also shown in patients with fibromyalgia. For example, compared to healthy controls, patients with fibromyalgia demonstrate greater MPFC activity during avoidance of punishment, and reduced MPFC activity during anticipation of reward (12,13). Similarly, midbrain activity is reduced in patients with fibromyalgia during periods of pain relief anticipation (i.e., rewarding event) and pain anticipation (i.e., punishment) (20). Likewise, the right striatum shows reduced response to reward in patients with chronic pain (mixed cohort of fibromyalgia or chronic low back pain) during anticipation of reward and loss (21).

Based on these data, we hypothesized that resting-state brain reward circuits (i.e., measured using fMRI-indicated functional connectivity) are altered in fibromyalgia, as in other types of chronic pain, such as low back pain. More specifically, we hypothesized that, similar to previous findings in patients with chronic back pain (during resting-state with rating task), NAcc-MPFC functional connectivity would be increased in patients with fibromyalgia.

Additionally, because previous resting-state studies in chronic back pain involved a pain-monitoring/rating task, we compared our results to a separate cohort of patients with chronic back pain during resting-state fMRI with no rating task. In our fibromyalgia cohort, we examined relationships between NAcc-MPFC functional connectivity and clinical measures of mood, affect, and symptom severity. Further, we conducted exploratory analyses of NAcc functional connectivity to mesolimbic (i.e., subcortical) circuit regions to clarify alterations in the mesolimbic valuation system in chronic pain.

## PARTICIPANTS AND METHODS

### Participants

Seventeen patients with fibromyalgia and 17 healthy individuals participated in the study. Patients with fibromyalgia were required to meet the modified American College of Rheumatology 2011 criteria for fibromyalgia: 1) a widespread pain index (WPI) score ≥ 7 + a symptom severity (SS) score ≥ 5, or WPI score 3-6 + SS score ≥ 9; 2) comparable symptoms present for at least 3 months; and 3) no diagnosis that would otherwise explain the pain (22). Other patient inclusion criteria were: pain in all 4 quadrants of the body, an average pain score of at least 2 (0-10 verbal scale) over the previous month, and no uncontrolled anxiety or depression. Because opioids can have dramatic and long-lasting effects on brain neurophysiology (23,24), all patients with fibromyalgia were relatively opioid naïve, in that they had not taken any opioid medications within 90 days prior to study participation, and their opioid usage was less than 1 month within their lifetime.

Healthy controls reported no history of chronic pain, took no pain medications at the time of the study visit, and did not have depression, anxiety, or major ongoing health conditions. Four patients reported taking no mood-altering or pain medications. The 13 remaining patients reported taking nonsteroidal anti-inflammatory drugs (NSAIDs, N = 7), gamma-aminobutyric acid (GABA) analogs (e.g., gabapentin and pregabalin, N = 6), serotonin-norepinephrine reuptake inhibitors (SNRIs, N = 4), tricyclic antidepressants (TCAs, N = 3), selective serotonin reuptake inhibitors (SSRIs, N = 2), low-dose naltrexone (LDN, N = 2), anticonvulsant drugs (N = 2), muscle relaxants (N = 2), other anxiolytics (e.g., buspirone hydrochloride, N=2), medical cannabis (N = 1), and using topical lidocaine patches (N = 1). We retained data from the participant taking medical cannabis in our final analysis as excluding this data did not significantly change the group results. Thirteen healthy controls were taking no mood-altering or pain medications. One healthy control participant took gabapentin (100 mg/day) and fluoxetine (40 mg/day) to treat premenstrual symptoms (2 days/month). Another healthy participant took celecoxib (200 mg) 3 weeks prior to study participation due to an ankle injury. We retained data from these 2 participants in our final analysis as excluding them did not significantly affect group results. All participants were required to have no MRI contraindications, and not be pregnant or nursing. All study procedures were approved by the Stanford University Institutional Review Board. A Data Use Authorization was created so that the data collected at Stanford University could be shared with/transferred to Duke University.

Prior to our analyses, we performed a power analysis to determine the sample size required to detect differences in NAcc-MPFC functional connectivity between patients with fibromyalgia and healthy controls. Figure 3b in Baliki et al. 2012 (11), which described functional connectivity (NAcc-MPFC) between patients with and without persistent pain, served as the input for our power analysis calculations. Results showed that we were well powered to detect group differences such that 17 participants per group would provide ≥ 80% power at alpha = 0.005 to detect a difference in mean connectivity of 0.05 between 2 groups of interest.

### Study Procedures

All participants were recruited at Stanford University, and data collection was conducted at the Richard M. Lucas Center for Imaging at Stanford University. Participants were first screened for any magnetic resonance contraindications. Then, they completed clinical questionnaires including the Beck Depression Inventory (BDI) (25), State-Trait Anxiety Inventory (STAI-State, STAI-Trait) (26), Behavioral Inhibition System/Behavioral Approach System Scales (BIS/BAS) (27), Profile of Mood States (POMS) (28), Positive and Negative Affect Schedule (PANAS) (29), Brief Pain Inventory – Short Form (BPI) (30), and PROMIS Fatigue (Item Bank v1.0, administered as computerized adaptive test) (31). After completing the questionnaires, MRI scans were conducted. Additional questionnaires and brain scans (12) were collected but were not included in the analyses for this study.

### MRI Scans

Neuroimaging data were acquired on a 3T General Electric scanner using an 8-channel head coil (GE Systems, Chicago, IL, USA). One T1 anatomic scan (3D FSPGR [fast spoiled gradient-echo] IRprep BRAVO) was performed for anatomic information. The image covered the whole brain, which included the brainstem and cerebellum, and parameters were as follows: 1-mm slice thickness, 22-mm frequency field of view (FOV), frequency direction anterior/posterior, number of excitations (NEX) 2, flip angle 11°, TR 6.8, TE 2.6, frequency 256, phase 256, and bandwidth 50.00. Functional scans consisted of a Gradient Echo Pulse Sequence using spiral in-out acquisition with 32 oblique slices acquired in a sequential descending slice order. The in-plane resolution was 2.0 mm x 2.0 mm, and slice thickness was 4.0 mm with 0.5-mm slice spacing (TR=2 seconds, TE = 30 seconds, flip angle 76°, pixel size 3.43 mm). By using the spiral in-out scan sequence, orbitofrontal signal drop-out was reduced (32), and acquisition of medial prefrontal and orbitofrontal cortices was improved. The resting-state scan consisted of 360 volumes.

### Image Preprocessing

Resting-state fMRI data were preprocessed using the default preprocessing pipeline in the CONN Toolbox v.20b (33) running in MATLAB v. R2020a and SPM12. Functional data underwent realignment and slice-timing correction. Outliers were identified based on the observed global blood oxygenation level-dependent (BOLD) fMRI signal and the amount of subject motion. The intermediate outlier identification setting in the CONN toolbox was used to identify framewise displacement (above 0.9 mm or global BOLD signal changes above 5 s.d.). T1-weighted structural images and functional images were segmented into grey matter (GM), white matter (WM), and cerebrospinal fluid (CSF) tissue classes, and normalized to Montreal Neurologic Institute (MNI) space. Lastly, functional data were smoothed using spatial convolution with a Gaussian kernel of 4 mm full width half maximum.

We then followed CONN’s default denoising pipeline (33). This step included an anatomic component-based noise correction procedure, corrections for potential confounding effects of WM and CSF, estimated subject-motion parameters, identified outlier scans or scrubbing, and bandpass filtering to 0.008-0.09 Hz. The number of outlier volumes detected ranged from 0% to 18.89% of the total volumes per participant. Rates were comparable to other published analyses (e.g., 34).

### ROI-to-VOI Analyses

Based on the approach used in a previous publication (11), 2 seed regions were defined: the right NAcc and the bilateral MPFC. The right NAcc region of interest (ROI, i.e., subcortical structure defined region) was created from the Desai Atlas included in Analysis of Functional NeuroImages (AFNI) (12) (Fig. 1A). The size of the bilateral MPFC volume of interest (VOI, i.e., defined by spherical volumes) was fixed at 10-mm diameter spheres centered at (−4, 50, -4; 4, 50, -4) (Fig. 1B). We used the CONN toolbox for the first-level analysis and group analysis. For first-level analyses, we used the ROI-to-VOI approach. We performed the first-level analysis to extract NAcc-MPFC functional connectivity per participant. Specifically, we computed a correlation analysis by averaging across the signal in each participant’s NAcc ROI and MPFC VOI, and correlating the signal of their right NAcc with their bilateral MPFC. We then performed group-level analyses such that correlation coefficients between the NAcc ROI and MPFC VOI for each participant were entered to identify differences in functional connectivity between groups. We also performed exploratory functional connectivity analyses by re-testing the primary hypothesis (i.e., functional connectivity between the right NAcc and bilateral MPFC) using the left NAcc and bilateral NAcc as alternative NAcc ROIs to measure functional connectivity with the bilateral MPFC.

**Figure 1.**
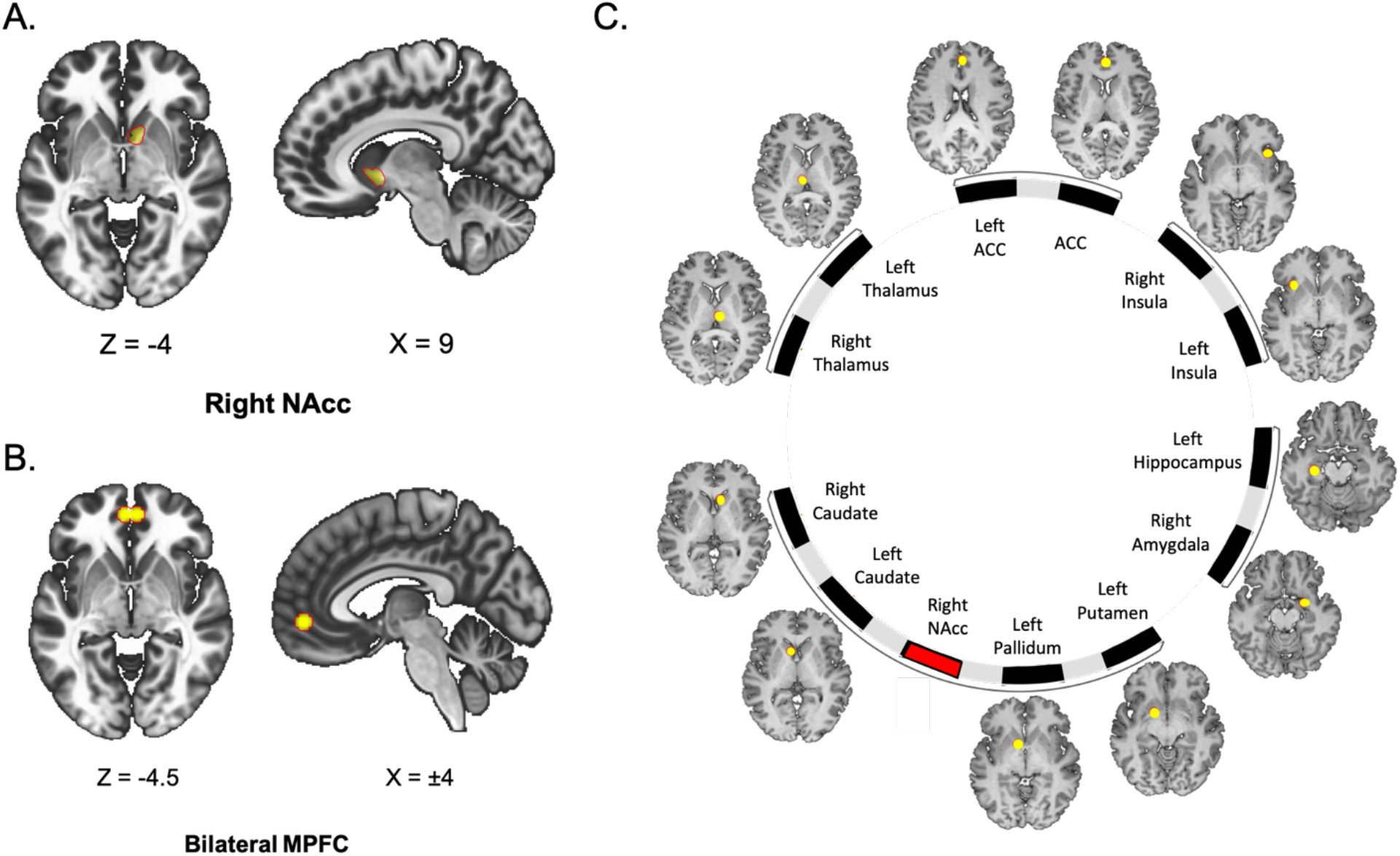
Regions and Volumes of Interest. (A) Location of the right NAcc ROI. (B) Location of bilateral MPFC VOI. (C) Location of 12 VOIs within mesolimbic circuits for exploratory analyses. Abbreviations: ROI, region of interest; VOI, volume of interest; NAcc, nucleus accumbens; MPFC, medial prefrontal cortex; ACC, anterior cingulate cortex.

To further characterize brain reward circuits in fibromyalgia, we investigated functional connectivity between other brain regions within mesolimbic circuits as additional exploratory VOI-based analyses as follows. We measured functional connectivity of the right NAcc with other regions within mesolimbic circuits including the hippocampus, amygdala, caudate, putamen, anterior cingulate cortex (ACC), insula, thalamus, and ventral pallidum (35). The volumes of interest were a fixed size of 10-mm diameter spheres centered at the left hippocampus (−30, -20, -18), right amygdala (24, -2, -16), left and right caudate (−8, 14, 2; 8, 20, 2), left putamen (−16, 4, -10), anterior cingulate cortex (2, 44, 20; 0, 44, 10), left and right insula (−32, 20, -4; 36, 20, -6), left and right thalamus (−6, -16, 8; 4, -14, 8), and left ventral pallidum (−10, 8, -4). The coordinates were determined based on previous findings of activation likelihood estimation and parametric voxel-based meta-analysis (36) (Fig. 1C).

### Connectivity vs. Clinical/Affective/Cognitive Measures Correlational Analyses

Correlational analyses between functional connectivity and questionnaire measures were performed within the patient group. Correlations between NAcc-MPFC functional connectivity and clinical-affective-cognitive data were tested using the general linear model in the CONN toolbox. Questionnaire data included clinical measures of pain duration and pain intensity (BPI), affect (PANAS), and fatigue (PROMIS Fatigue). Fisher-transformed correlation coefficient values from the first-level analysis were tested for correlations with questionnaire data. Additional exploratory correlational analyses between NAcc-MPFC connectivity and depression (BDI), anxiety (State-Trait Anxiety), reward-related behavioral inhibition/activation (BIS/BAS Subscales), and total mood disturbance (POMS) were also performed.

### Post-hoc Comparison to Chronic Back Pain Cohort

We conducted resting-state fMRI, which did not involve any task during scan; however, there were methodologic differences between our study and the previous study (10,11), such that prior research (10,11) analyzed functional connectivity from fMRI scans with thermal stimulation or a continuous self-report pain-monitoring task. Thus, we compared our NAcc-MPFC functional connectivity results from patients with fibromyalgia to a separate cohort of patients with chronic back pain who underwent a similar condition of resting-state fMRI (i.e., no pain rating task was involved).

For the chronic back pain dataset, we analyzed fMRI scan data from the “cbp_resting” project provided by the Open Pain Project (OPP) database (principal investigator: A. Vania Apkarian; http://www.openpain.org; all participants in this dataset consented to participate in the study approved by the Institutional Review Board at Northwestern University.) The dataset contains 34 patients with chronic back pain, but we excluded 3 participants after preprocessing because their scans could not be aligned.

The same right NAcc ROI and bilateral MPFC VOI described above were used for NAcc-MPFC functional connectivity analysis of this comparison dataset. Data were preprocessed using the default preprocessing pipeline in the CONN Toolbox v.20b (33) running in MATLAB v. R2020a and SPM12. Functional data underwent realignment, but slice-timing correction was not performed. Outliers were identified from the observed global BOLD fMRI signal and the amount of subject motion. The intermediate outlier identification setting in the CONN Toolbox was used to identify framewise displacement. T1-weighted structural images and functional images were segmented into GM, WM, and CSF classes, and normalized to MNI space. Lastly, functional data were smoothed using spatial convolution with a Gaussian kernel of 4 mm full width half maximum. Next, CONN’s default denoising pipeline was followed (33). The number of outlier volumes detected ranged from 0.1% to 14% of the total volumes per participant.

For functional connectivity analysis, the same 2 seed regions that were defined in our main analysis (i.e., NAcc-MPFC functional connectivity) were used: the right NAcc ROI and the bilateral MPFC VOI (Fig. 1A). As we did in our main analysis, we performed a correlation analysis by averaging across the signal in each of the ROIs and VOIs, and correlating the signal of the right NAcc with the bilateral MPFC for the first-level analysis. Next, correlation coefficients between the ROI and VOI for each subject were entered to identify differences in connectivity between groups. CONN Toolbox was used for these first- and second-level analyses. We again conducted additional exploratory VOI-based analyses, such that we measured functional connectivity of the right NAcc with 12 regions within mesolimbic circuits. The 12 VOIs that were defined in our exploratory analyses (Fig. 1C) were used.

### Statistical Analysis

For determining group differences in our primary endpoint of NAcc-MPFC functional connectivity, we used an unpaired two-sample *t*-test of correlation coefficients, thresholded for significance at *P* < 0.05. For functional connectivity-questionnaire measures correlational analyses, the standard *P* < 0.05 was used to evaluate the significance. For the additional analyses of regional functional connectivity (i.e., exploratory analyses), we similarly used unpaired two-sample *t*-tests of correlation coefficients, thresholded for significance at *P* < 0.05. Correction for multiple comparisons was not applied for the exploratory analyses.

## RESULTS

### Participant Demographics, Medications, and Clinical Measures

Data from 17 fibromyalgia patients and 17 healthy controls were included in the analysis. (See Table 1 for participant demographics.) Duration of pain related to fibromyalgia symptoms ranged from 2-28 years (M = 11.5 years, SD = 7.7 years). Clinical/affective/cognitive measures, including mood disturbance, fatigue, anxiety, depression, pain distribution across the body, pain severity, and pain interference, were significantly higher in patients compared to controls (Table 2). Patients’ widespread pain symptoms across the body is presented in Fig. 2.

**Table 1.** Participants’ Demographic Information: One patient was left-handed, and one healthy control did not report handedness. “Other” race refers to other than Asian, Caucasian, African American, Hispanic or Latina, Pacific Islander or Alaskan, or Native American. One patient and 2 healthy controls did not report their income level. Education levels are divided into “High School” (up to or through high school), “College/University” (up to or through college/university), and “Advanced Degrees” (post college/university).

|  | Patients | Controls |
| --- | --- | --- |
| **Total Participants (all female)** | **17** | **17** |
| **Righthanded** | **16** | **16** |
| **Self-Identified Race** |  |  |
| **Asian** | **2** | **7** |
| **Caucasian** | **13** | **9** |
| **Other** | **2** | **1** |
| **Hispanic or Latina Ethnicity** | **3** | **1** |
| **Employment Status** |  |  |
| **Part-time employed** | **3** | **2** |
| **Full-time employed** | **6** | **11** |
| **Unemployed** | **8** | **4** |
| **Income Level** |  |  |
| **\$0–\$29,999** | **5** | **0** |
| **\$30,000–\$59,999** | **3** | **2** |
| **\$60,000 or more** | **8** | **13** |
| **Education Level** |  |  |
| **High School** | **3** | **0** |
| **College/University** | **11** | **8** |
| **Advanced Degree** | **3** | **9** |

**Table 2.** Participant Demographics, Medications, and Clinical Measures. Due to incomplete questionnaires, the number of participants for each measure differs from the total participant counts. *P* values shown are not corrected for multiple comparisons. Abbreviations: sd, standard deviation.

|  | Patients |  | Controls |  | P-Value |
| --- | --- | --- | --- | --- | --- |
|  | N | Mean±sd | N | Mean±sd |  |
| Age | 17 | 48.1±9.6 | 17 | 48.4±10.3 | 0.997 |
| Positive Affect (PANAS) | 17 | 26.4±8.5 | 17 | 35.7±5.5 | 0.0017 |
| Negative Affect (PANAS) | 17 | 21.2±8.0 | 17 | 13.1±3.8 | 0.0019 |
| Behavioral Drive (BAS) | 16 | 13±4.8 | 16 | 13.1±5.2 | 0.953 |
| Behavioral Fun (BAS) | 16 | 12.4±4.1 | 16 | 13.8±4.3 | 0.265 |
| Behavioral Reward (BAS) | 16 | 19.1±5.5 | 16 | 19.3±5.7 | 0.831 |
| Behavioral Inhibition (BIS) | 16 | 26.5±7.6 | 16 | 20.4±7.6 | < 0.001 |
| Total Mood Disturbance (POMS) | 17 | 21.6±15.7 | 17 | -4.4±8 | < 0.001 |
| Fatigue (PROMIS) | 17 | 65.5±8.0 | 17 | 49±6.8 | < 0.001 |
| Trait Anxiety (STAI) | 17 | 49.7±8.5 | 17 | 34.5±7.8 | < 0.001 |
| State Anxiety (STAI) | 17 | 41.4±7.0 | 17 | 26.9±7 | < 0.001 |
| Depression (BDI) | 17 | 15.8±8.9 | 16 | 2.2±2.8 | < 0.001 |
| Number of Pain Areas (FAF) | 17 | 13.9±3.9 | 17 | 1.5±1.8 | < 0.001 |
| Pain Severity (BPI) | 17 | 5.7±2.1 | 17 | 0.4±1.0 | < 0.001 |
| Pain Interference (BPI) | 17 | 5.3±2.7 | 17 | 0.5±0.9 | < 0.001 |

**Figure 2.**
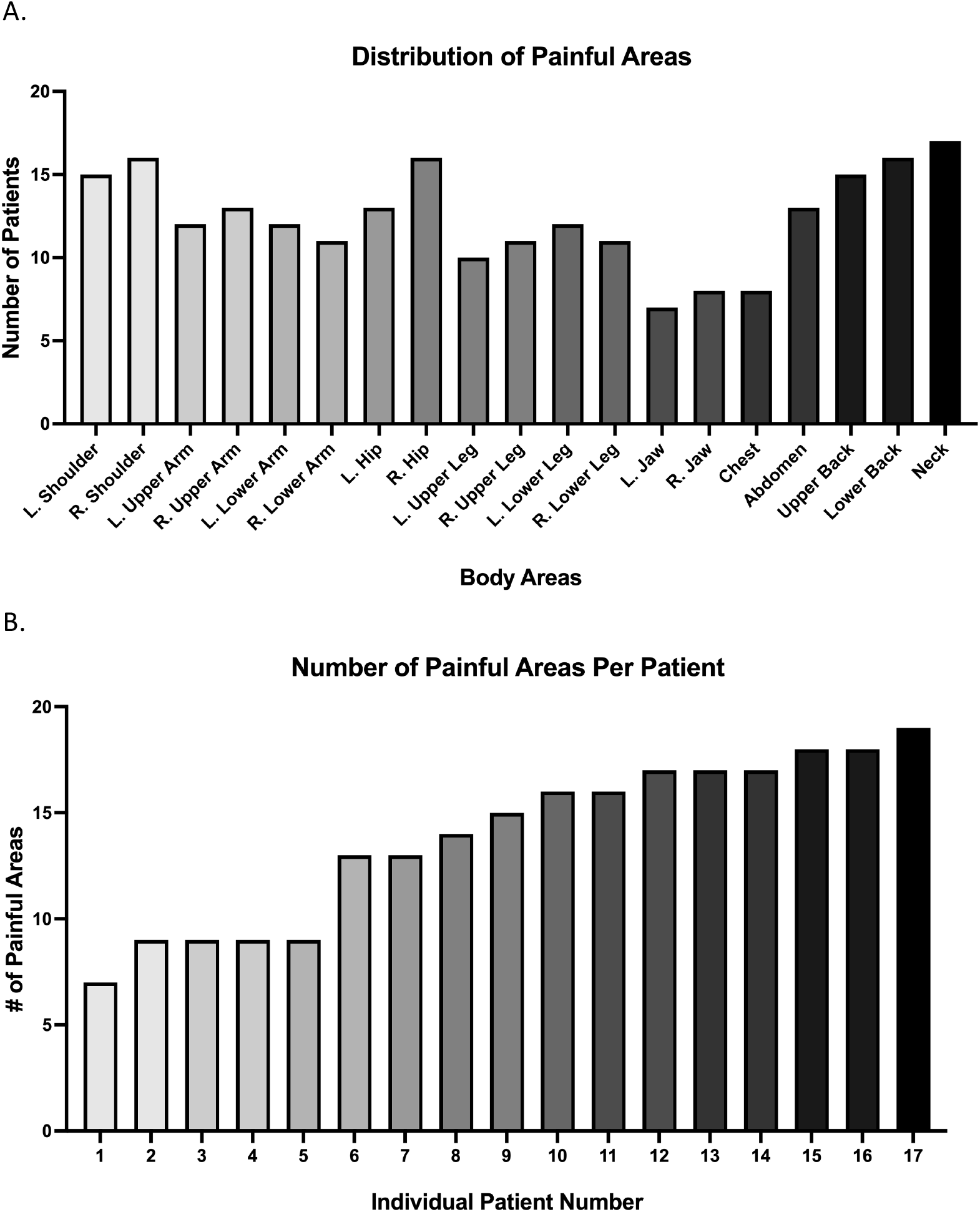
Distribution and Number of Painful Areas in Patients. (A) Painful body areas are shown across19 body regions (listed in the Fibromyalgia Assessment Form; x-axis). The presence (or absence) of pain in each body region was reported by each patient (total N = 17 patients; y-axis). (B) Each patient is represented by a number, and patients are ordered by low to high number of painful body areas (x-axis). The number of painful body areas reported by each patient is shown (total painful areas N = 19; y-axis). In both figures, different bar shading is for data readability, and does not represent additional results. Abbreviations: L., left; R., right.

### NAcc-MPFC Circuit Functional Connectivity

The main analysis and our primary endpoint focused on group differences in NAcc-MPFC functional connectivity (Fig. 3A). Robust NAcc-MPFC functional connectivity was found among patients with fibromyalgia and controls (t(16) = 4.2, *P* = 0.0006 and t(16) = 4.71, *P* = 0.0002, respectively), and no significant group differences were observed (P = 0.323; Fig. 3B).

**Figure 3.**
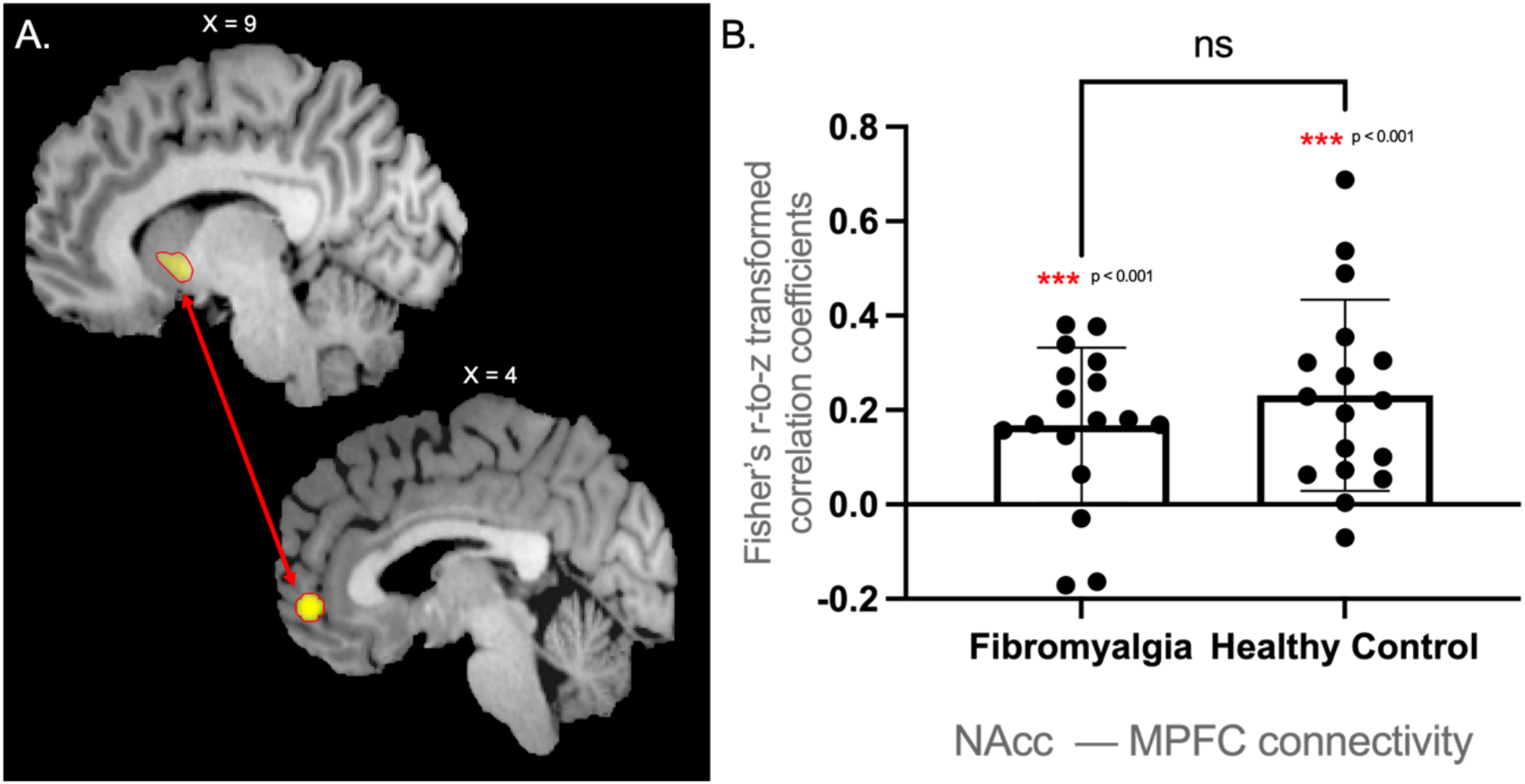
NAcc-MPFC Functional Connectivity in Patients with Fibromyalgia vs. Healthy Controls. (A) Right NAcc ROI (top) – Bilateral MPFC VOI (bottom). (B) NAcc-MPFC functional connectivity in patients with fibromyalgia and healthy controls. Dots represent data points for individual participants. Abbreviations: ROI, region of interest; VOI, volume of interest; NAcc, nucleus accumbens; MPFC, medial prefrontal cortex; ns, not significant (color should be used in print).

The right NAcc region of interest (ROI) and bilateral MPFC volume of interest (VOI) were predetermined based on the approach used in Martucci et al. 2018 (12). As both this study and our present study have strong predetermined hypotheses (i.e., primary analyses restricted to the NAcc and MPFC), we followed the previous approach in creating the right NAcc ROI and bilateral MPFC VOI. To determine whether a slightly different ROI may lead to different patterns of results, we also created the same ROI used by Baliki et al. 2012 (11) (i.e., 5-mm radius spheres centered at [10, 12, -8] for the right NAcc ROI and at [2, 52, -2] for the right MPFC ROI) as an exploratory analysis. We followed the same statistical analysis steps (i.e., image preprocessing and ROI-to-VOI analysis) used for our primary analysis. Again, the results confirmed robust NAcc-MPPFC functional connectivity in both groups (patients with fibromyalgia and healthy controls, t(16) = 5.76, P = 0.00003 and t(16) = 5.65, P = 0.00003, respectively), and no significant group differences (P = 0.762).

### Correlations with Clinical, Affective, and Cognitive Measures

Within the patient group, correlational analysis of functional connectivity vs. questionnaire measures did not show a significant correlation between NAcc-MPFC functional connectivity and pain duration (*P* = 0.3). Further, no significant correlations were identified between NAcc-MPFC functional connectivity and clinical pain intensity, affect, and fatigue (all *P* > 0.1). For exploratory analyses, we assessed the extent to which NAcc-MPFC connectivity correlated with other clinical, affective, and cognitive measures. We found only a positive correlation trend of increased NAcc-MPFC functional connectivity with total mood disturbance (Table 3).

**Table 3.** Correlations between fMRI and Questionnaire Measures in Patients. Tested relationships between NAcc-MPFC circuit functional connectivity and clinical, affective, cognitive measures are shown. The analyses were exploratory, and *P* values shown were not corrected for multiple comparisons.

|  | **P-value**<br>**(Correlation with NAcc – MPFC**<br>**functional connectivity)** |
| --- | --- |
| **Positive Affect (PANAS)** | 0.7159 |
| **Negative Affect (PANAS)** | 0.1831 |
| **Behavioral Reward (BAS)** | 0.101 |
| **Behavioral Drive (BAS)** | 0.3596 |
| **Behavioral Fun (BAS)** | 0.2568 |
| **Behavioral Inhibition (BIS)** | 0.0864 |
| **Total Mood Disturbance (POMS)** | 0.0635 |
| **Fatigue (PROMIS)** | 0.3625 |
| **Trait Anxiety (STAI)** | 0.1519 |
| **State Anxiety (STAI)** | 0.5478 |
| **Depression (BDI)** | 0.469 |
| **Pain Severity (BPI)** | 0.234 |
| **Pain Interference (BPI)** | 0.1927 |

### NAcc-Mesolimbic Circuit Functional Connectivity

For exploratory analyses, we re-tested the primary analysis question (i.e., functional connectivity between the right NAcc and bilateral MPFC) using the left NAcc and bilateral NAcc. These served as alternative NAcc ROIs to compare functional connectivity with the bilateral MPFC in patient vs. healthy control groups. No significant differences were observed between groups (all *P* > 0.1). We then measured functional connectivity of the right NAcc with 12 regions within mesolimbic circuits, including the ACC, left hippocampus, left ACC, left caudate, left putamen, left insula, left thalamus, left ventral pallidum, right amygdala, right caudate, right insula, and right thalamus. Compared to controls, patients with fibromyalgia showed a trend of decreased connectivity of the right NAcc with ACC (t(32) = -1.91, *P* = 0.065) and significantly reduced connectivity with the left putamen (t(32) = -2.11, *P* = 0.0428), right thalamus (t(32) = -2.11, *P* = 0.0423), and left ventral pallidum (t(32) = -3.62, *P* = 0.0009) (all uncorrected; Fig. 4A and 4B). Connectivity values between all 13 regions (including the right NAcc, with each region as a seed) are shown in Figure 4C.

**Figure 4.**
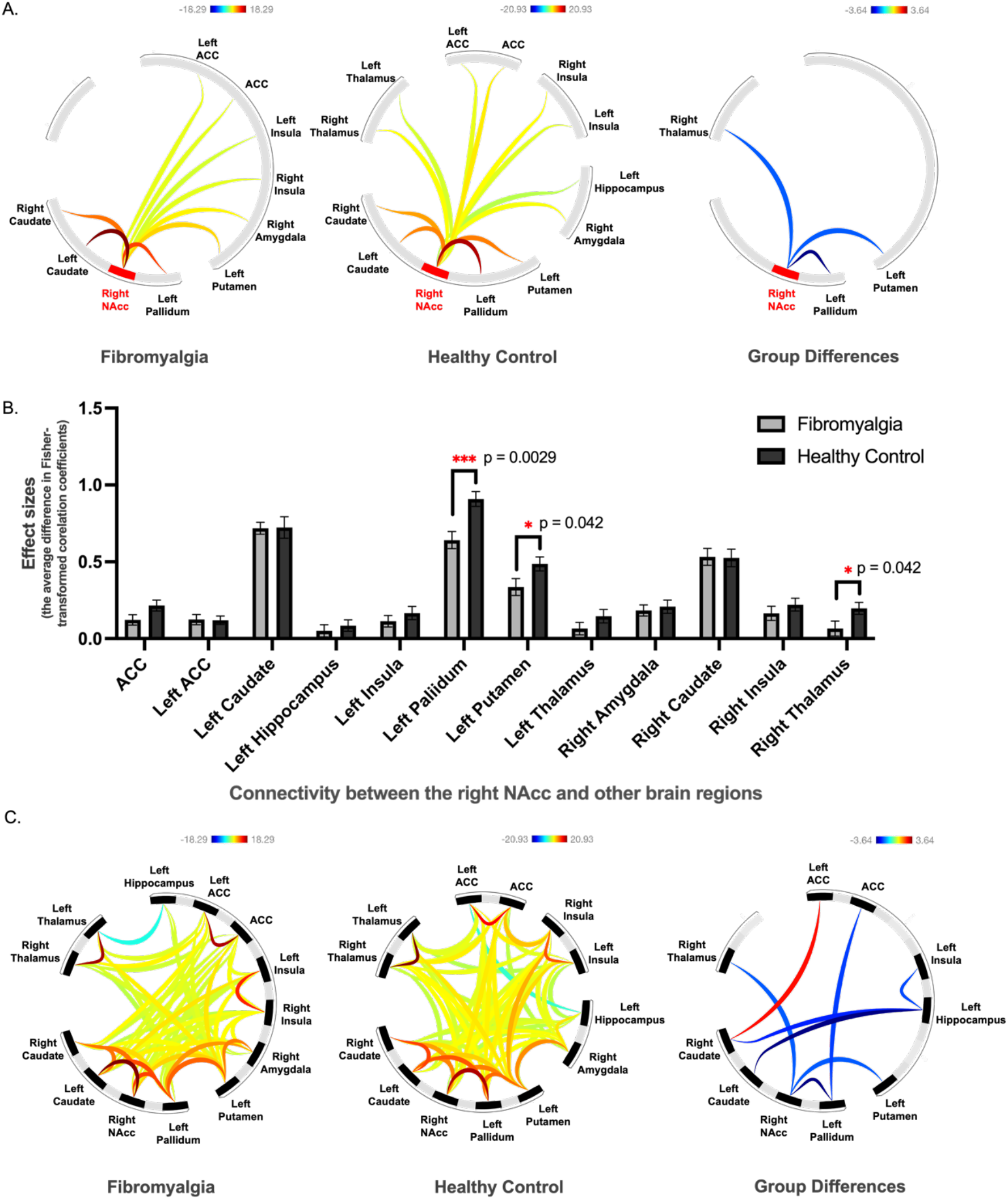
Functional Connectivity Values between the Right NAcc and 12 Brain Regions within Mesolimbic Circuits. (A) From left to right, the figures represent functional connectivity in the patient group, healthy control group, and group differences. For the group difference, the contrast used was Fibromyalgia > Healthy controls (e.g., reduced left putamen - right NAcc connectivity in patients with fibromyalgia compared to healthy controls). (B) The average difference in Fisher-transformed correlation coefficients (i.e., effect sizes) in 12 brain regions in each group. (C) All functional connectivity values between 13 brain regions (including the right NAcc). From left to right, the figures represent functional connectivity in the patient group, healthy control group, and group differences. For the group difference, the contrast used was Fibromyalgia > Healthy controls. In figures A and C, line color indicates the magnitude of the effect for ROI-to-VOI functional connectivity. Blue - light blue color lines indicate negative connectivity, and yellow - red lines indicate positive connectivity. Results are thresholded for significance at *P* < 0.05 (uncorrected for multiple comparisons). Abbreviations: ROI, region of interest; VOI, volume of interest; NAcc, nucleus accumbens; ACC, anterior cingulate cortex (color should be used in print).

### NAcc-MPFC Circuit Connectivity in a Separate Patient Cohort

Unlike previous findings in patients with low back pain (10,11), no differences in NAcc-MPFC functional connectivity between patients with fibromyalgia and controls were observed. Several possibilities may account for this difference, including the possibility that brain reward system alterations may differ between various chronic pain types. Further, there were methodologic differences between our study and the previous study (10,11). Therefore, we compared our NAcc-MPFC functional connectivity results from patients with fibromyalgia to a separate cohort of patients with chronic back pain who underwent similar resting-state fMRI, which did not involve any task during scan.

Robust NAcc-MPFC functional connectivity was found in both patient groups (fibromyalgia: t(16) = 4.198, *P* < 0.001; chronic back pain: t(30) = 5.948, *P* < 0.001), but no significant group differences were observed (*P* > 0.9; two-sample *t*-test, assuming unequal variance; Fig. 5). We also measured the average difference in Fisher-transformed correlation coefficients (i.e., effect sizes) between the right NAcc and 12 brain regions in mesolimbic circuits in patients with fibromyalgia, patients with chronic back pain, and healthy controls. These results are shown in Figure 6.

**Figure 5.**
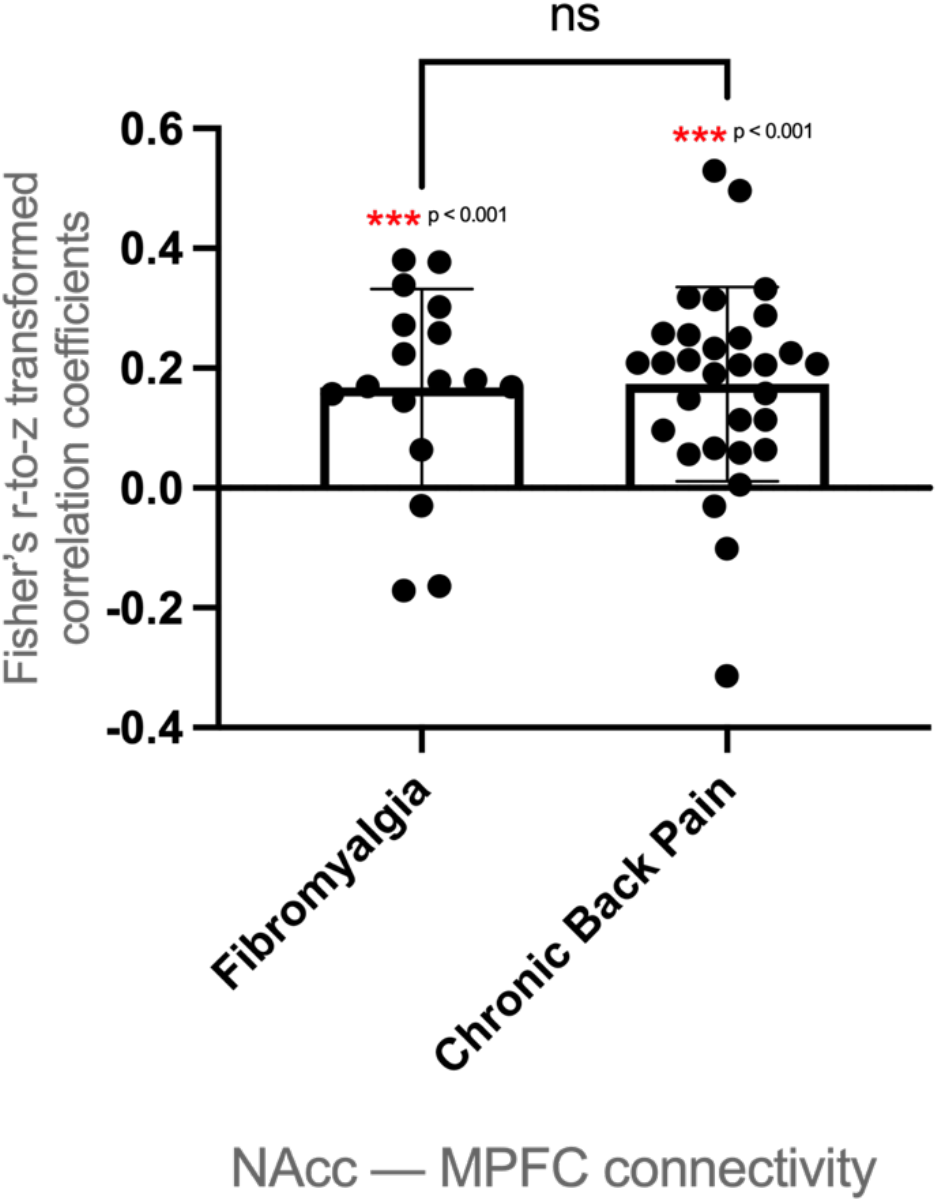
Comparison to a Separate Cohort Data Set. NAcc-MPFC functional connectivity in patients with fibromyalgia and a separate cohort of patients with chronic back pain from the OPP database (openpain.org). Dots represent data points for individual participants. Abbreviations: OPP, Open Pain Project; NAcc, nucleus accumbens; MPFC, medial prefrontal cortex; ns, not significant.

**Figure 6.**
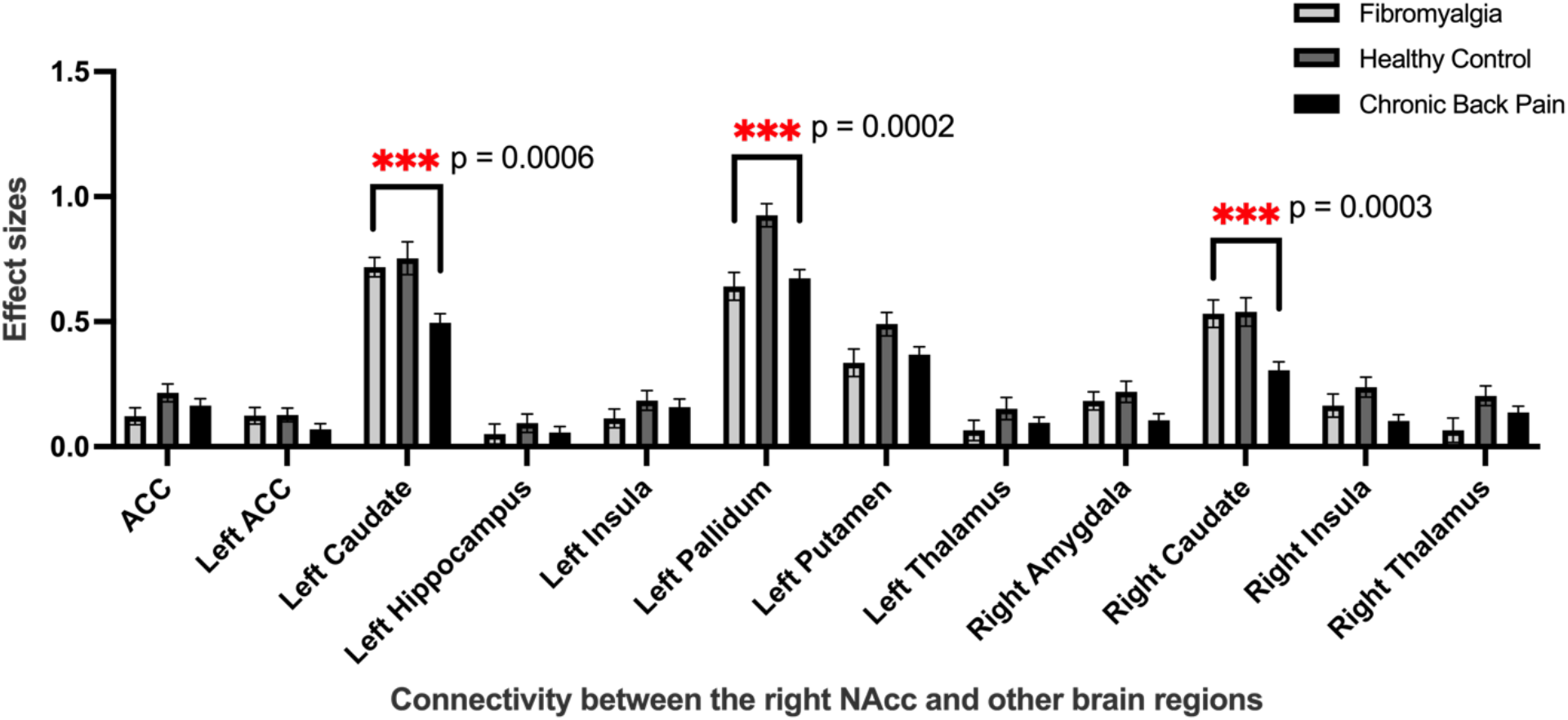
Average Difference in Fisher-transformed Correlation Coefficients (i.e., effect sizes) in Patients with Fibromyalgia vs. Chronic Back Pain vs. Healthy Controls. One-way ANOVA for each region revealed significant differences in left and right caudate (F(2, 62) = 8.36, *P* = 0.0006, F (2, 62) = 9.1, *P* = 0.0003), and left pallidum (F (2, 62) = 9.3, *P* = 0.0002). The right NAcc-caudate (left and right) functional connectivity was significantly reduced in patients with chronic back pain compared to fibromyalgia and controls (all *P* < 0.001, uncorrected); and, similar to patients with fibromyalgia, patients with chronic back pain showed reduced right NAcc-left pallidum connectivity compared to healthy controls (all *P* < 0.001, uncorrected). Abbreviations: ACC, anterior cingulate cortex; NAcc, nucleus accumbens.

## DISCUSSION

Based on previous findings (10,11), we hypothesized that patients with fibromyalgia would exhibit greater resting-state fMRI-indicated functional connectivity of the NAcc-MPFC reward circuit compared to healthy controls. However, while robust NAcc-MPFC functional connectivity was identified in both patients with fibromyalgia and healthy controls, no group differences were observed during resting-state. Similarly, we found no group differences when comparing resting-state NAcc-MPFC functional connectivity in patients with fibromyalgia to a separate cohort of patients with chronic back pain (resting-state fMRI dataset from openpain.org). Correlational analysis of functional connectivity and questionnaire measures in patients with fibromyalgia showed only a trend toward a significant association between greater NAcc-MPFC functional connectivity and total mood disturbance.

Extending NAcc functional connectivity analysis to the broader mesolimbic reward circuits revealed novel evidence of mesolimbic reward/valuation system alterations in fibromyalgia vs. controls. Specifically, patients with fibromyalgia demonstrated significantly reduced functional connectivity of the right NAcc with the left putamen, right thalamus, and left ventral pallidum. Overall, these results suggest that corticostriatal circuit (i.e., NAcc-MPFC) functional connectivity during the resting state condition is intact, yet may be impacted by variable cognitive demands and experiences (i.e., resting-state vs. task-involved fMRI scans). Meanwhile, our data indicate that other NAcc-mesolimbic circuits are altered in fibromyalgia during the resting state, which complements prior observations of altered brain reward response to behavioral cues (e.g., 12,13) and implicates reward-system/circuit wide alterations in patients with fibromyalgia.

### Intact NAcc-MPFC Circuit Resting-State Functional Connectivity in Fibromyalgia

We present the first investigation of brain reward circuits, specifically functional connectivity of the key NAcc-MPFC circuit and broader NAcc-mesolimbic functional connectivity, in fibromyalgia. This study contributes to prior neuroimaging research that has identified altered brain reward activity in patients with fibromyalgia (12,13,20,37) and provides insight into directions for future research on brain reward circuits (i.e., functional connectivity) in chronic pain.

Several factors may impact the differences between the previous findings (i.e., increased NAcc-MPFC functional connectivity in patients with chronic back pain) (10,11) and our results. First, demographic differences (e.g., sex distribution, our study: all female, Baliki et al. (2010): 8 males and 8 females) and/or different clinical status (e.g., pain duration, our study: mean = 11.5 years, Baliki et al. (2010): mean = 7.86 years) may have differently influenced our results compared to previous published findings. Second, while administration of noxious thermal stimuli or a self-report pain-rating task were used during fMRI scans in previous functional connectivity based studies (10,11), no task and/or stimulation experience was involved during acquisition of our resting-state fMRI data in our fibromyalgia cohort (and chronic back pain cohort included here). These methodologic differences may impact the results, such that pain stimulation (and coinciding cognitive evaluation/experience of painful stimuli) may drive increased NAcc-MPFC functional connectivity in chronic pain conditions vs. controls (10). A continuous self-report pain-monitoring task was also involved when researchers found greater NAcc-MPFC functional connectivity in patients with sub-acute back pain (SBP) whose pain persisted vs. patients with SBP who recovered (11). This finding lends further support to a hypothesis that pain stimulation as well as intensity of ongoing chronic pain (and coinciding cognitive evaluation/experience) influence NAcc-MPFC functional connectivity. Several studies have investigated brain circuits in fibromyalgia during task-based states (e.g., 38). Our present conflicting results underscore the notion that these nuanced differences (i.e., chronic pain type/subgroups; task vs. resting-state conditions) are important to consider in future evaluations of brain reward circuits in chronic pain.

To evaluate our follow-up hypothesis that mesolimbic functional connectivity alterations vary by chronic pain type, we compared NAcc-MPFC functional connectivity in our fibromyalgia cohort to a cohort of patients with chronic back pain. Importantly, these data from chronic back pain patients were collected in a resting-state status when no task was involved, thereby controlling for this potential methodologic influence on the results. Surprisingly, from this analysis, no significant differences in functional connectivity were observed between patients with fibromyalgia and chronic back pain. This null result is not sufficient to deduce that NAcc-MPFC functional connectivity is generalizable between chronic pain types; it must be evaluated prospectively across matched cohorts for different chronic pain types to determine generalizability (e.g., different MRI parameters due to the nature of using an openly available data set for comparison here). Still, it does suggest that use of a pain stimulus/pain monitoring task during fMRI scans may substantially influence NAcc-MPFC functional connectivity in patients with chronic back pain.

### Altered NAcc-Mesolimbic Circuit Functional Connectivity in Fibromyalgia

Within the broader mesolimbic system, analysis of NAcc circuits in patients with fibromyalgia revealed significantly reduced functional connectivity between the NAcc and the left ventral pallidum, left putamen, and right thalamus. All of these brain regions play essential roles in reward processing. Specifically, the ventral pallidum is a critical region in reward and motivational processing, and encodes positive reward signals (39). The left putamen is activated during anticipation of reward, and increased left putamen activity is associated with positive reward (40). Similarly, the right thalamus is activated during periods of increasing reward (41) and during receipt of both reward and punishment outcomes (42). In addition, previous functional connectivity studies have revealed a critical role for thalamic projections in shaping and supporting reward processing within the NAcc among both healthy adults and adolescents (43). Reduced functional connectivity between the putamen and NAcc has been demonstrated in patients with anorexia nervosa (44), a condition highly related to brain reward system alterations (45,46). Similar to these prior studies in other healthy and clinical populations, our findings of reduced functional connectivity within mesolimbic circuits lend further support to the clinical relevance of reward system alterations among patients with fibromyalgia. Indeed, changes in functional connectivity predict and track with treatment response in fibromyalgia (47,48), which underscores the value of evaluating brain reward circuit alterations, in chronic pain.

### Correlations with Clinical, Affective, and Cognitive Measures

Several previous studies in patients with fibromyalgia have identified relationships between altered resting-state fMRI activity and clinical status and affective measures (49–51). To our knowledge, no prior studies have specifically examined potential relationships between NAcc-MPFC functional connectivity and various clinical, affective, and cognitive measures in fibromyalgia. Correlational analyses between NAcc-MPFC functional connectivity and our included questionnaire measures largely did not reveal any significant relationships (e.g., pain duration, pain intensity, affect, fatigue). Notably, however, we found a positive, although not significant, correlational trend between increased NAcc-MPFC functional connectivity and total mood disturbance. While we were not able to observe greater NAcc-MPFC functional connectivity in patients with fibromyalgia compared to healthy controls, this positive correlational trend demonstrates that patient variability (from low to high NAcc-MPFC functional connectivity) may still relate to mood disturbance. Indeed, previous studies have shown that striatal dysfunction is highly related to mood disorders (52), such that striatal hypofunction and pain-comorbid mood alternation occur together (21). Our results complement these findings, and provide preliminary data for a potential relationship between reward system alterations and mood disturbance in chronic pain.

### Limitations and Future Directions

Previous research reported NAcc-MPFC functional connectivity as a biomarker of pain chronification (11) in patients with chronic back pain, therefore we initially hypothesized that NAcc-MPFC functional connectivity would be positively correlated with pain duration within patient groups. Because we did not identify significantly altered NAcc-MPFC functional connectivity in our fibromyalgia cohort, it follows that we were not able to identify any significant relationships between NAcc-MPFC functional connectivity and our questionnaire measures, including pain duration. However, our results indicate the possibility that NAcc-MPFC functional connectivity is not a biomarker of pain chronification in fibromyalgia, despite its relevance in chronic back pain. In addition, for the power analysis, we primarily focused on the group difference of NAcc-MPFC functional connectivity between patients and controls, while the connectivity-clinical correlation was analyzed only within the fibromyalgia group. Thus, the lack of identified correlation between connectivity and our questionnaire measures may derive from a lack of power for the correlation analyses.

Our findings of altered NAcc connectivity to other regions within mesolimbic reward circuits in fibromyalgia were from exploratory analyses, and must be evaluated prospectively in future investigations. The patterns of altered functional connectivity observed between all 13 regions (Figure 6) generate numerous possibilities regarding reward system alterations in fibromyalgia, and it is essential that future studies examine the reproducibility of these findings in patients with fibromyalgia prospectively via testable hypotheses. Ultimately, future investigations must link functional connectivity between brain regions with specific roles in reward processing (e.g., reward/punishment anticipation, reward/punishment outcomes), and assess relationships between reward circuit regions during resting-state vs. task-based states as well as consider effects of medications. Further, future studies aimed to determine how altered reward circuits may influence and interact with other components of the central nervous system that are altered in fibromyalgia (53) will help to clarify the complex neurobiology of fibromyalgia and chronic pain more generally.

## Conclusion

Our study examined resting-state functional connectivity in reward circuits among patients with fibromyalgia compared with controls, based on hypotheses generated from previous findings of reward circuit alterations in patients with chronic back pain. The absence of significant group differences in NAcc-MPFC functional connectivity during resting-state fMRI condition provides evidence supporting intact corticostriatal circuit functional connectivity in fibromyalgia. A comparison between our results and a separate dataset of patients with chronic back pain suggests putative impacts of variable cognitive demands and experiences on altered corticostriatal circuit connectivity in chronic pain. Our exploratory analyses underscore the presence of potentially widespread mesolimbic reward/valuation system alterations in fibromyalgia, and the need for future research to more fully understand the complex neural mechanisms associated with fibromyalgia.

## Data Availability

The data that support the findings of this study are available on request from the corresponding author, KTM. The data are not publicly available due to their containing information that could compromise the privacy of research participants.

## Acknowledgment

This work was pre-registered on the Open Science Framework (OSF, https://osf.io/cj9u8). This project was funded by the National Institutes of Health, National Institute of Drug Abuse (NIDA), K99/R00 DA040154 (awarded to KTM). Data used in this project from the Open Pain Project (OPP; Principal Investigator: A. Vania Apkarian, PhD, Northwestern University) was funded by the National Institute of Neurological Disorders and Stroke (NINDS) and National Institute of Drug Abuse (NIDA). The authors thank Kathy Gage for very helpful editorial comments on the manuscript.

## Notes

Conflict of Interest Statement: The authors have no conflicts of interest to disclose.

### Competing Interest Statement

The authors have declared no competing interest.

### Author Declarations

All study procedures were approved by the Stanford University Institutional Review Board. A Data Use Authorization was created so that the data collected at Stanford University could be shared with/transferred to Duke University. All participants in this dataset consented to participate in the study approved by the Institutional Review Board at Northwestern University (principal investigator: A. Vania Apkarian; http://www.openpain.org).

## REFERENCES

1. Fields H. A motivation-decision model of pain: The role of opioids. Proc 11th World Congr Pain 2006:449–459.

2. Becerra L, Borsook D. Signal valence in the nucleus accumbens to pain onset and offset. Eur J Pain Lond Engl 2008;12:866–869.

3. Becerra L, Breiter HC, Wise R, Gonzalez RG, Borsook D. Reward circuitry activation by noxious thermal stimuli. Neuron 2001;32:927–946.

4. Borsook D, Linnman C, Faria V, Strassman AM, Becerra L, Elman I. Reward deficiency and anti-reward in pain chronification. Neurosci Biobehav Rev 2016;68:282–297.

5. DosSantos MF, Moura B de S, DaSilva AF. Reward Circuitry Plasticity in Pain Perception and Modulation. Front Pharmacol 2017;8:790.

6. Gatchel RJ, Peng YB, Peters ML, Fuchs PN, Turk DC. The biopsychosocial approach to chronic pain: scientific advances and future directions. Psychol Bull 2007;133:581–624.

7. Kamping S, Bomba IC, Kanske P, Diesch E, Flor H. Deficient modulation of pain by a positive emotional context in fibromyalgia patients. PAIN® 2013;154:1846–1855.

8. Nees F, Ruttorf M, Fuchs X, Rance M, Beyer N. Brain-behaviour correlates of habitual motivation in chronic back pain. Sci Rep 2020;10:11090.

9. Berger SE, Baria AT, Baliki MN, Mansour A, Herrmann KM, Torbey S, et al. Risky monetary behavior in chronic back pain is associated with altered modular connectivity of the nucleus accumbens. BMC Res Notes 2014;7:739.

10. Baliki MN, Geha PY, Fields HL, Apkarian AV. Predicting value of pain and analgesia: nucleus accumbens response to noxious stimuli changes in the presence of chronic pain. Neuron 2010;66:149–160.

11. Baliki MN, Petre B, Torbey S, Herrmann KM, Huang L, Schnitzer TJ, et al. Corticostriatal functional connectivity predicts transition to chronic back pain. Nat Neurosci 2012;15:1117–1119.

12. Martucci KT, Borg N, MacNiven KH, Knutson B, Mackey SC. Altered prefrontal correlates of monetary anticipation and outcome in chronic pain. Pain 2018;159:1494–1507.

13. Martucci KT, MacNiven KH, Borg N, Knutson B, Mackey SC. Apparent Effects of Opioid Use on Neural Responses to Reward in Chronic Pain. Sci Rep 2019;9:9633.

14. Haber SN, Calzavara R. The cortico-basal ganglia integrative network: the role of the thalamus. Brain Res Bull 2009;78:69–74.

15. Haber SN, Knutson B. The reward circuit: linking primate anatomy and human imaging. Neuropsychopharmacol Off Publ Am Coll Neuropsychopharmacol 2010;35:4–26.

16. Sescousse G, Caldú X, Segura B, Dreher J-C. Processing of primary and secondary rewards: a quantitative meta-analysis and review of human functional neuroimaging studies. Neurosci Biobehav Rev 2013;37:681–696.

17. Knutson B, Adams CM, Fong GW, Hommer D. Anticipation of increasing monetary reward selectively recruits nucleus accumbens. J Neurosci Off J Soc Neurosci 2001;21:RC159.

18. Camara E, Rodriguez-Fornells A, Ye Z, Münte TF. Reward networks in the brain as captured by connectivity measures. Front Neurosci 2009;3:350–362.

19. Tzschentke TM. The medial prefrontal cortex as a part of the brain reward system. Amino Acids 2000;19:211–219.

20. Loggia ML, Berna C, Kim J, Cahalan CM, Gollub RL, Wasan AD, et al. Disrupted brain circuitry for pain-related reward/punishment in fibromyalgia. Arthritis Rheumatol Hoboken NJ 2014;66:203–212.

21. Kim M, Mawla I, Albrecht DS, Admon R, Torrado-Carvajal A, Bergan C, et al. Striatal hypofunction as a neural correlate of mood alterations in chronic pain patients. NeuroImage 2020;211:116656.

22. Wolfe F, Clauw DJ, Fitzcharles M-A, Goldenberg DL, Häuser W, Katz RS, et al. Fibromyalgia criteria and severity scales for clinical and epidemiological studies: a modification of the ACR Preliminary Diagnostic Criteria for Fibromyalgia. J Rheumatol 2011;38:1113–1122.

23. Upadhyay J, Maleki N, Potter J, Elman I, Rudrauf D, Knudsen J, et al. Alterations in brain structure and functional connectivity in prescription opioid-dependent patients. Brain 2010;133:2098–2114.

24. Younger JW, Chu LF, D’Arcy NT, Trott KE, Jastrzab LE, Mackey SC. Prescription opioid analgesics rapidly change the human brain. PAIN 2011;152:1803–1810.

25. Beck AT, Steer RA, Carbin MG. Psychometric properties of the Beck Depression Inventory: Twenty-five years of evaluation. Clin Psychol Rev 1988;8:77–100.

26. Spielberger CD, Gorsuch RL, Lushene RE. Manual for the State-Trait Anxiety Inventory. 1970. Available at: http://ubir.buffalo.edu/xmlui/handle/10477/2895. Accessed July 16, 2021.

27. Carver CS, White TL. Behavioral inhibition, behavioral activation, and affective responses to impending reward and punishment: The BIS/BAS Scales. J Pers Soc Psychol 19950101;67:319.

28. McNair DM, Lorr M, Droppleman LF. Manual profile of mood states. 1971.

29. Watson D, Clark LA, Tellegen A. Development and validation of brief measures of positive and negative affect: the PANAS scales. J Pers Soc Psychol 1988;54:1063–1070.

30. Keller S, Bann CM, Dodd SL, Schein J, Mendoza TR, Cleeland CS. Validity of the brief pain inventory for use in documenting the outcomes of patients with noncancer pain. Clin J Pain 2004;20:309–318.

31. Cella D, Riley W, Stone A, Rothrock N, Reeve B, Yount S, et al. The Patient-Reported Outcomes Measurement Information System (PROMIS) developed and tested its first wave of adult self-reported health outcome item banks: 2005-2008. J Clin Epidemiol 2010;63:1179–1194.

32. Glover GH, Law CS. Spiral-in/out BOLD fMRI for increased SNR and reduced susceptibility artifacts. Magn Reson Med 2001;46:515–522.

33. Whitfield-Gabrieli S, Nieto-Castanon A. Conn: a functional connectivity toolbox for correlated and anticorrelated brain networks. Brain Connect 2012;2:125–141.

34. Koh J, Kaneoke Y, Donishi T, Ishida T, Sakata M, Hiwatani Y, et al. Increased large-scale inter-network connectivity in relation to impulsivity in Parkinson’s disease. Sci Rep 2020;10:11418.

35. Richard JM, Castro DC, Difeliceantonio AG, Robinson MJF, Berridge KC. Mapping brain circuits of reward and motivation: in the footsteps of Ann Kelley. Neurosci Biobehav Rev 2013;37:1919–1931.

36. Liu X, Hairston J, Schrier M, Fan J. Common and distinct networks underlying reward valence and processing stages: a meta-analysis of functional neuroimaging studies. Neurosci Biobehav Rev 2011;35:1219–1236.

37. Jensen KB, Srinivasan P, Spaeth R, Tan Y, Kosek E, Petzke F, et al. Overlapping structural and functional brain changes in patients with long-term exposure to fibromyalgia pain. Arthritis Rheum 2013;65:3293–3303.

38. Jarrahi B, Martucci KT, Nilakantan AS, Mackey S. Cold Water Pressor Test Differentially Modulates Functional Network Connectivity in Fibromyalgia Patients Compared with Healthy Controls. Annu Int Conf IEEE Eng Med Biol Soc IEEE Eng Med Biol Soc Annu Int Conf 2018;2018:578–582.

39. Smith KS, Tindell AJ, Aldridge JW, Berridge KC. Ventral pallidum roles in reward and motivation. Behav Brain Res 2009;196:155–167.

40. Knutson B, Fong GW, Adams CM, Varner JL, Hommer D. Dissociation of reward anticipation and outcome with event-related fMRI. Neuroreport 2001;12:3683–3687.

41. Elliott R, Friston KJ, Dolan RJ. Dissociable neural responses in human reward systems. J Neurosci Off J Soc Neurosci 2000;20:6159–6165.

42. Knutson B, Westdorp A, Kaiser E, Hommer D. FMRI visualization of brain activity during a monetary incentive delay task. NeuroImage 2000;12:20–27.

43. Cho YT, Fromm S, Guyer AE, Detloff A, Pine DS, Fudge JL, et al. Nucleus accumbens, thalamus and insula connectivity during incentive anticipation in typical adults and adolescents. NeuroImage 2013;66:508–521.

44. Favaro A, Tenconi E, Degortes D, Manara R, Santonastaso P. Effects of obstetric complications on volume and functional connectivity of striatum in anorexia nervosa patients. Int J Eat Disord 2014;47:686–695.

45. Wagner A, Aizenstein H, Venkatraman VK, Fudge J, May JC, Mazurkewicz L, et al. Altered reward processing in women recovered from anorexia nervosa. Am J Psychiatry 2007;164:1842–1849.

46. Bischoff-Grethe A, McCurdy D, Grenesko-Stevens E, Irvine LEZ, Wagner A, Yau W-YW, et al. Altered brain response to reward and punishment in adolescents with Anorexia nervosa. Psychiatry Res 2013;214:331–340.

47. Schmidt-Wilcke T, Ichesco E, Hampson JP, Kairys A, Peltier S, Harte S, et al. Resting state connectivity correlates with drug and placebo response in fibromyalgia patients. NeuroImage Clin 2014;6:252–261.

48. Cummiford CM, Nascimento TD, Foerster BR, Clauw DJ, Zubieta J-K, Harris RE, et al. Changes in resting state functional connectivity after repetitive transcranial direct current stimulation applied to motor cortex in fibromyalgia patients. Arthritis Res Ther 2016;18:40.

49. Napadow V, LaCount L, Park K, As-Sanie S, Clauw DJ, Harris RE. Intrinsic brain connectivity in fibromyalgia is associated with chronic pain intensity. Arthritis Rheum 2010;62:2545–2555.

50. Napadow V, Kim J, Clauw DJ, Harris RE. Decreased intrinsic brain connectivity is associated with reduced clinical pain in fibromyalgia. Arthritis Rheum 2012;64:2398–2403.

51. Čeko M, Frangos E, Gracely J, Richards E, Wang B, Schweinhardt P, et al. Default mode network changes in fibromyalgia patients are largely dependent on current clinical pain. NeuroImage 2020;216:116877.

52. Borsook D, Becerra L, Carlezon WA, Shaw M, Renshaw P, Elman I, et al. Reward-aversion circuitry in analgesia and pain: implications for psychiatric disorders. Eur J Pain Lond Engl 2007;11:7–20.

53. Martucci KT, Weber KA, Mackey SC. Altered Cervical Spinal Cord Resting-State Activity in Fibromyalgia. Arthritis Rheumatol Hoboken NJ 2019;71:441–450.

